# The COVID19 pandemic has changed women’s experiences of pregnancy in the UK

**DOI:** 10.1101/2021.11.05.21265698

**Authors:** Sarah Sturrock, Kim Turner, Chelone Lee-Wo, Vanessa Greening, Asma Khalil, Paul T Heath, Kirsty Le Doare

## Abstract

**Introduction:** During the SARS-CoV-2 pandemic, maternity care has been substantially altered to reduce transmission of the SARS-CoV-2 virus. Many antenatal services are now restricted or delivered online, and visiting has been restricted during labour and in the postnatal period.

**Methods:** We conducted an online survey from 1^st^ August to 31^st^ December 2020 to investigate the experiences of women who were pregnant or breastfeeding in the UK during the SARS-CoV-2 pandemic. The survey included 55 open and closed questions and required 5 minutes to complete. We publicised the survey using social media.

**Results:** We received 96 responses, including 66 currently pregnant women and 22 women who were pregnant during the pandemic. The response rate was 70.1% of survey views. We found mixed experiences of the impact of the pandemic on antenatal and perinatal care, notably with some women feeling visiting restrictions were insufficient and others feeling they were too strict. Twenty-nine women received no information about COVID-19, and 6 women found it very difficult to find information. Thirty-nine women would have liked to have more information about breastfeeding after a pregnancy affected by COVID-19, and 37 women wanted more information about antibody persistence and transfer.

**Discussion:** Additional support is required for pregnant and lactating women during the current pandemic. Provision of information and support, including via social media, may improve women’s experiences of pregnancy in the current environment.

**Significance:** Maternity services in the UK have been significantly restructured to prevent transmission of the SARS-CoV-2 virus, including restrictions to in-person antenatal care, and perinatal visiting. It is not fully known how these changes are perceived by pregnant and breastfeeding women.

Reactions to changes in antenatal care are mixed, including whether restrictions were too lenient or too strict. Most women underwent online antenatal care in addition in-person visits. Some received no information about COVID-19, and a significant proportion of women would have liked more information, particularly regarding antibody transfer and benefits of breastfeeding during the pandemic.

**Ethical statement:** This study was approved by North East - Newcastle & North Tyneside 1 Research Ethics Committee

## Introduction

To limit the transmission of COVID-19, substantial changes in the provision of maternity care have been made worldwide, particularly in high income countries. In the first months of the pandemic, guidelines which support maternity health services changed constantly, as models of healthcare during pregnancy were adapted to meet physical distancing requirements.(Khalil et al. 2021) Many antenatal care services were restricted or offered online only the UK. Many hospitals introduced a visitor restriction policy for labour and birth, and some allowed only a partner to be physically present in the hospital.(Brown 2020)(Royal College of Obstetricians and Gynaecologists 2020)

## Methods

We undertook to understand the experiences of women who were planning a pregnancy, were currently pregnant or were breastfeeding in the UK during the SARS-CoV-2 pandemic using an online survey designed using the SurveyMonkey® tool, promoted online using social media between 1^st^ August to 31^st^ December 2020. There were 55 open and closed questions within the survey and women were informed they would require approximately 5 minutes to complete. Information was provided on data storage and use, Investigator name and the purpose of the study. The data was tested internally before launching via social media to check for internal validity and logic, then disseminated via the periCOVID Facebook page and Twitter. The periCOVID website is designed to provide information on pregnancy and the neonatal period during the pandemic (www.pericovid.com). Women were asked to complete the survey directly into the tool by following the link. No participant identifiable information was collected. A validity check was built into the survey such that the survey responses could not be submitted if certain questions were not answered. Respondents were able to review and change their answers before they were submitted. Because of the nature of the survey we were unable to check for multiple entries per individual. Data was analysed according to emerging themes and displayed as number (%). We describe the survey using the CHERRIES checklist.(Eysenbach 2004)

## Results

Ninety-six women completed the survey, of whom 66 were currently and 22 had recently been pregnant (Table 1). We had 120 views on the Facebook page and 17 retweets. If each click and retweet is considered one individual viewing the questionnaire then our response rate was 70.1%.

**Table 1.** Summary of antenatal care experiences.

| Type of care |  | Number of women (%) | Comments |
| --- | --- | --- | --- |
| **Antenatal care visits** | N=96 |  |  |
|  | In person | 28 (29%) |  |
|  | Online and in person | 66 (69%) |  |
|  | Online only | 2 (2%) |  |
| **Routine blood pressure and urine monitoring** | N=66 |  |  |
|  | In clinic | 52 (83%) |  |
|  | At home | 8 (12%) | One woman did not know what to do if results abnormal |
| **Hospital contact** | N=62 |  |  |
|  | Midwife number if concerned | 59 (90%) |  |
| **Complications of pregnancy** | N=66 |  |  |
|  | None | 36 (56%) |  |
|  | High blood pressure | 6% |  |
|  | Gestational diabetes | 6% |  |
|  | SARS-COV-2 | 5 (%) |  |
|  | Stillbirth | 1 |  |
|  | Ectopic pregnancy | 1 |  |
|  | Admission antenatally | 8 |  |
| **SARS-COV-2 Testing** |  |  |  |
|  | Yes | 25 |  |
|  | No | 37 |  |
| **Testing reason** |  |  |  |
|  | Close contact | 18% (4) |  |
|  | Symptoms of COVID19 | 45% (10) | Most experienced mild symptoms and remained at home whilst one had moderate symptoms and was admitted to hospital. |
|  | Routine testing of all women | 36% (8) |  |
| **Delivery care** |  |  |  |
|  | Virtual tour of the delivery suite | 14 (22%) |  |
|  | In person tour of the delivery suite | 38 (61%) |  |
|  | Partner, friend of family attendance in labour | 40 (62%) |  |
| Postnatal care | N= 28 |  |  |
|  | No visitors allowed | 7 (11%) |  |
|  | Visitor hours restricted | 21 (33%) |  |
|  | Only partners allowed | 10 (16%) |  |
| Information about COVID19 | N=47 |  |  |
|  | Easy to understand | 15 (62%) |  |
|  | very difficult to find more information, | 6 (4%) |  |
|  | neither easy or difficult to find information. | 26 (44%) |  |
|  | None received | 50% (29) |  |
| Source of information accessed | RCOG | 61% (35) |  |
|  | Tommy's | 54% (31) |  |
|  | PHE | 42% (24) |  |
|  | Midwife | 29% (17) |  |
|  | Government briefings | 21% (12) |  |
| Information on |  |  |  |
|  | Reducing the risk of SARS-CoV-2 infection to themselves during pregnancy | 62% (37) | Of those who had received information the majority found it straightforward to understand and act on. |
|  | Vertical transmission | 87% (58) |  |
|  | Reducing the risk of transmission during labour and delivery | 81% (45) |  |
|  | Reducing the risk of infection once their baby had been born | 90% (50) |  |
|  | Reducing the risk of infection whilst breastfeeding | 92% (50) |  |

When asked if women felt that their birth choices were limited by the pandemic 15 either strongly agreed or agree, whilst 17 disagreed or strongly disagreed, and 16 felt that the pandemic and reorganisation of maternity services had no impact on their choices. Two women were not able to have their partners with them for their first scan. One woman who gave birth five days before the first national lockdown was disappointed not to be able to use the birthing pool in her unit due to staff shortages. One woman who was asked to shield during the pandemic did not feel reassured by staff that she and her baby would be protected from infection during their time in hospital. Reactions to postnatal visiting restrictions were also mixed; one woman felt that there should have been more restrictions as she didn’t feel protected and safe. Three women found the uncertainty regarding the rules unsettling and would have liked better guidance from the hospital.

Thirty-nine women responded that they would like to have more information on the benefits of breastfeeding after a pregnancy affected by COVID-19 disease; 37 of women expressed an interest in having more information on how long the antibodies that pregnant women produce when they are infected persist, if these antibodies cross the placenta to the baby and how long they persist in both the mother and the baby after delivery. Regarding vaccination against COVID-19, 46% (24) women felt that pregnant women should not be included in vaccine trials (Figure 1).

**Figure 1.**
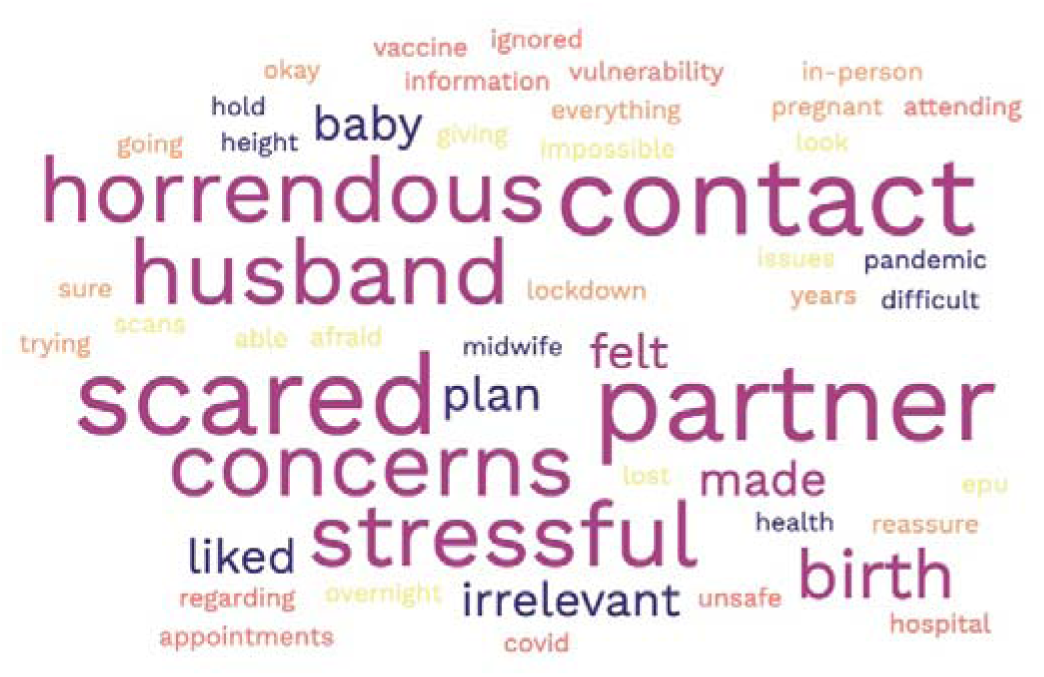
Word cloud of common themes in comments from pregnant and lactating women

## Discussion

Special support is required for pregnant women yet there are key information gaps that pregnant and breastfeeding women would like to access to improve their experiences of pregnancy in the pandemic. Social media sites linked directly to maternity services are vital to engage women, as these emerged as primary sources of support and information.

## Data Availability

All data produced in the present study are available upon reasonable request to the authors

## Acknowledgements

We would like to thank the pregnant women who took part in this survey.

